# Social networks and social support during cervical cancer treatment among women at a tertiary referral facility in Ghana: A qualitative descriptive study

**DOI:** 10.64898/2026.10.05.26364081

**Authors:** Fawzia Rufai, Rebecca Essah Dowuona, Doreen Annung-yen, Dickson Kweku Delali, Awolu Adam

## Abstract

**Background:** Cervical cancer remains a leading cause of cancer death among women in sub-Saharan Africa, with Ghana experiencing persistently high mortality linked to late diagnosis and constrained treatment access. Beyond its clinical burden, cervical cancer treatment is a socially demanding experience requiring women to mobilise financial, practical, informational, and emotional support over time. This study explored the social networks surrounding women during cervical cancer treatment and the social support exchanged within them at a tertiary referral facility in Ghana.

**Methods:** A qualitative descriptive design, informed by an interpretivist epistemological orientation, was employed. Eight women receiving treatment at the National Centre for Radiotherapy, Oncology and Nuclear Medicine, Korle-Bu Teaching Hospital, Accra, were recruited using criterion-based purposive sampling and interviewed individually using a semi-structured guide until data saturation was reached. Interviews were audio-recorded, transcribed verbatim, and analysed using reflexive thematic analysis, supported by Atlas.ti version 7.5.7.

**Results:** Five themes were generated. Disclosure functioned as a gatekeeping process through which women selectively determined who entered their support network, shaped by anticipated stigma. Despite valuing the support they received, women displayed marked reluctance to request it directly, relying on indirect strategies and expressing gratitude regardless of whether support met their actual needs. Support networks consistently narrowed around a single family member, most often a daughter or sibling, who provided the bulk of financial, practical, and informational assistance, while more peripheral relationships receded. Social support functioned not merely as emotional comfort but as a precondition for accessing and continuing treatment at all, particularly given widely inadequate insurance coverage. Finally, some women experienced discomfort reconciling their need for support with a prior identity as habitual providers of care to others.

**Conclusion:** Social networks and support during cervical cancer treatment in Ghana are actively constructed through disclosure, constrained by reluctance to seek help, and concentrated around a single caregiver, functioning as a determinant of whether treatment can be accessed and sustained rather than merely a source of comfort. Formalizing support around, rather than in place of, these informal networks could strengthen both access to and continuity of cervical cancer care in similarly resourced settings.

## Background

Cervical cancer remains a major public health challenge worldwide, despite being largely preventable through Human papillomavirus (HPV) vaccination, screening, and timely treatment [1]. In 2022, an estimated 662,301 new cases and 348,874 deaths occurred globally, making cervical cancer the fourth most common cancer and fourth leading cause of cancer death among women [1]. The burden is unequally distributed, with markedly higher incidence and mortality in countries with lower levels of development [2]. If current rates persist, the global burden is projected to rise to 760,082 cases and 411,035 deaths by 2030 [1].

Sub-Saharan Africa bears the highest cervical cancer burden globally and remains the region where the disease causes the greatest mortality among women [3,4]. In 2020, the region recorded about 110,300 cervical cancer cases, and cervical cancer was the leading cause of female cancer death in 27 countries [5]. More recent estimates show that in 2023 sub-Saharan Africa had an age-standardized incidence of 55.9 per 100,000 and mortality of 27.1 per 100,000, more than two-to four-fold global rates [3]. The burden is also heterogeneous within the region, with Eastern sub-Saharan Africa carrying the highest rates and Western sub-Saharan Africa the lowest [6].

African trends are concerning because the burden is not declining in parallel with global progress [7]. Across the African Union, age-standardized incidence rose from 17.41 to 23.53 per 100,000 between 1990 and 2023 [7]. Southern Africa has shown the steepest long-term increases, while all sub-Saharan African subregions showed a post-2020 uptick [3]. Without substantial improvements in prevention and care, incidence and mortality are projected to more than double in many African countries by 2050 [8].

These patterns reflect persistent gaps in HPV vaccination, screening, and treatment access, compounded in some settings by high HIV prevalence and weak health-system capacity [3,8]. Across low-resource settings, cervical cancer outcomes are strongly shaped by late diagnosis and limited access to definitive treatment [9]. Mortality in sub-Saharan Africa remains high, with pooled estimates ranging from about 32% in one meta-analysis to 56% in another, reflecting differences in study design and follow-up but consistently poor survival [9,10]. Advanced stage at diagnosis is the strongest and most consistent predictor of death in the region [10].

Ghana sits within this broader West African context of high burden, constrained treatment access, and poor outcomes. Although Western sub-Saharan Africa has lower rates than Eastern and Southern Africa, its 2023 burden remained substantial at 35.7 incident cases and 17.8 deaths per 100,000 women [3]. Ghana has introduced national cancer control strategies and a nationwide HPV vaccination campaign, indicating policy recognition of cervical cancer as a major public health concern [11]. Even so, Ghanaian outcome data included in regional mortality syntheses show persistently high mortality among women diagnosed with cervical cancer [9].

Within sub-Saharan African meta-analytic evidence, Ghana-specific studies contributed to a pooled mortality estimate of 0.66, placing the country among the higher-burden settings represented in survival analyses [9]. Poorer outcomes in Ghana are linked to the same factors seen elsewhere in the region, especially advanced-stage disease, incomplete treatment, and restricted access to comprehensive care [9]. These patterns mean that many women begin treatment with complex clinical, financial, and practical needs that extend beyond the biomedical management of disease [12].

The social dimensions of treatment become especially important in this context. Cervical cancer disproportionately affects women in midlife in Africa, with a concentrated burden among women aged roughly 40–59 years, when family and economic responsibilities are often greatest [7,8]. Treatment in low-resource settings often unfolds amid transport barriers, delayed referrals, treatment interruptions, and weak continuity of care [6]. Under such conditions, women’s ability to start, continue, and complete treatment depends not only on clinical services but also on whether they can mobilize financial, practical, informational, emotional, and spiritual support through their social networks [13].

That is the key justification for the present study. Epidemiologic research clearly defines the scale and inequity of cervical cancer in sub-Saharan Africa and Ghana, but it says much less about how women actually navigate treatment through everyday relationships [5]. A qualitative study focused on women’s social networks during cervical cancer treatment can therefore add an important missing layer by showing who becomes involved, how disclosure shapes that involvement, what forms of support are exchanged, and whether women perceive that support as adequate to meet treatment-related needs, including their ability to access and continue treatment itself. In that sense, the study contributes not by re-establishing that cervical cancer is a major burden, but by explaining how women live through treatment within that burden.

## Methods

### Study design

This study employed a qualitative descriptive design, informed by an interpretivist epistemological orientation, to explore how women experience and describe their social networks and the social support exchanged within them during cervical cancer treatment, including which relationships became involved, what forms of support were exchanged, and how women perceived the usefulness and adequacy of that support in relation to their treatment and wellbeing. A qualitative descriptive approach was considered appropriate given that women’s social network and support experiences during cervical cancer treatment remain a relatively understudied phenomenon in this context, requiring rich, contextual description that remains close to participants’ own accounts, rather than theory generation or in-depth phenomenological interpretation [14,15]. Reporting of the study followed the Consolidated Criteria for Reporting Qualitative Research (COREQ) to ensure transparent and comprehensive presentation of the study context, methods, analysis, and findings [16].

### Study setting

This study was conducted at the National Centre for Radiotherapy, Oncology and Nuclear Medicine, located within the Korle-Bu Teaching Hospital in Accra, Ghana. Korle-Bu Teaching Hospital, established in 1923, is one of the largest teaching hospitals in Africa and serves as a major tertiary referral centre for Ghana and the wider West African sub-region [17]. The National Centre for Radiotherapy, Oncology and Nuclear Medicine, operational since 1997, is one of the largest cancer treatment facilities in the sub-region, offering radiotherapy (external beam and brachytherapy), chemotherapy, and other specialist oncology services to paediatric and adult patients with solid tumours [18]. As a tertiary referral centre, it receives women with cervical cancer requiring specialist treatment from Accra and other parts of Ghana, making it an appropriate setting for exploring their social network and support experiences during treatment.

### Study population

The study population comprised women receiving treatment for cervical cancer at the National Centre for Radiotherapy, Oncology and Nuclear Medicine, Korle-Bu Teaching Hospital.

### Inclusion and exclusion criteria

Participants were eligible for inclusion if they were adult women with a confirmed diagnosis of cervical cancer, currently receiving treatment at the National Centre for Radiotherapy, Oncology and Nuclear Medicine, Korle-Bu Teaching Hospital, and able to provide informed consent. Women were excluded if they were too physically or psychologically unwell to participate in an interview, unable to provide informed consent due to cognitive impairment, or receiving palliative care only with no active treatment at the centre.

### Sampling and sample size

A criterion-based purposive sampling technique was used to recruit women who met the study’s eligibility criteria and could provide firsthand, information-rich accounts relevant to the study objectives. This approach was considered appropriate for a qualitative descriptive design, as it prioritizes depth and relevance of participant experience over statistical representativeness [14]. Sample size was guided by the principle of data saturation, whereby recruitment continued until no substantively new codes or patterns emerged from successive interviews; saturation was judged to have been reached after eight interviews. This is broadly consistent with evidence from a systematic review of empirical saturation studies, which found that qualitative research with relatively homogeneous populations and narrowly defined objectives commonly reaches saturation within nine to seventeen interviews, with smaller samples more plausible where study scope is tightly bounded, as in the present single-site, treatment-phase-specific sample [19].

Fifteen women were initially approached for participation. Of these, seven did not participate: three were unable to take part because they felt too physically weak or unwell following treatment, two were scheduled for interviews on three separate occasions but were unable to attend, and two declined because they preferred to keep their cervical cancer diagnosis private. Eight women ultimately participated in the study.

### Pilot interviews

Prior to data collection, the interview guide and probes were reviewed by members of the research team, a senior research fellow, and personnel from the Cancer Registry office at Korle-Bu Teaching Hospital to identify potential problems and assess the cultural sensitivity of the questions. The interview procedures and guide, including the introductory questionnaire, interview questions, and probes, were subsequently piloted with two women who were not included in the final study sample. Findings from the pilot informed refinements to the interview procedures and questions prior to the main data collection.

### Data collection instrument and procedure

Data were collected using a semi-structured interview guide developed by the research team to elicit detailed accounts of participants’ social networks and the support they received during cervical cancer treatment. The guide was organized around five broad areas: socio-demographic and clinical background; cervical cancer treatment history; the composition of participants’ social networks and the support exchanged within them; access to healthcare resources; and the perceived impact of social support on treatment adherence, quality of life, and self-perceived wellbeing. Open-ended questions were accompanied by probes to encourage elaboration and allow unanticipated issues raised by participants to be explored in depth. The guide was refined following expert review and piloting, as described above.

Interviews were conducted face-to-face by a member of the research team/the principal investigator, in a private space within/near the National Centre for Radiotherapy, Oncology and Nuclear Medicine, to ensure privacy and minimize disruption to participants’ care. Interviews were conducted in English/Twi, with interpretation where necessary and lasted approximately 45-75 minutes. All interviews were audio-recorded with participants’ consent and supplemented with brief field notes capturing contextual observations and non-verbal cues. Data collection took place between August 16 and September 14, 2024. Recruitment and interviewing continued until data saturation was reached, as described in the sampling section above.

Audio recordings were transcribed verbatim by the research team, and transcripts were checked against the original recordings for accuracy before analysis. Transcripts were translated into English where interviews were conducted in a local language, with attention to preserving participants’ original meaning. Identifying information was removed during transcription, and each participant was assigned a unique alphanumeric code to preserve anonymity throughout analysis and reporting.

## Data analysis

Audio recordings were transcribed verbatim, and interviews conducted in local dialects were translated into English. Transcripts were checked against the original recordings and field notes to confirm accuracy and to preserve contextual and non-verbal information where relevant.

Data were managed and coded using Atlas.ti version 7.5.7 and analysed using reflexive thematic analysis. Transcripts were read repeatedly to achieve familiarity with the dataset before coding began. Initial codes were then generated inductively and semantically, remaining close to participants’ own language rather than to the structure of the interview guide. Related codes were subsequently clustered into candidate subthemes, which were reviewed against the coded data and the full dataset to confirm that each was coherent, distinct, and adequately supported. Candidate subthemes were then organised into overarching themes capturing patterns that recurred across, rather than within, individual transcripts. This process was iterative rather than linear: earlier coding decisions were revisited and, where necessary, revised as later transcripts were analysed and as candidate themes were tested against the full dataset. Coding was guided by the study objectives while remaining open to concepts that emerged inductively from participants’ accounts beyond the original interview framework. Disconfirming cases, where a participant’s account diverged from the dominant pattern within a theme, were retained and reported rather than treated as noise.

Analytic memos and reflective notes were maintained throughout to document coding decisions, track the development of themes, and record instances where later analysis prompted revision of earlier interpretations. Dependability and confirmability were supported through peer debriefing, in which the research supervisor/a co-investigator reviewed and challenged emerging codes and themes at key points in the analysis, and through an audit trail documenting coding decisions, data management steps, and analytic memos. As all data were generated through audio-recorded, semi-structured individual interviews, with transcripts verified against recordings and field notes for completeness, missing data did not apply to this study.

### Trustworthiness

The trustworthiness of this study was addressed using the criteria of credibility, dependability, confirmability, and transferability.

### Credibility

It was supported through several strategies. Purposive sampling was used to recruit women who could provide detailed, firsthand accounts relevant to the study objectives, increasing the likelihood that the data reflected genuine and information-rich experiences. The interview guide was reviewed by members of the research team, a senior research fellow, and personnel from the Cancer Registry office prior to data collection, and subsequently piloted with two women outside the final sample, allowing the instrument to be refined before use. All quotations presented in the findings were verified against the verbatim transcripts at the point of writing to guard against misattribution or drift from participants’ original wording. Where participants’ accounts diverged from the dominant pattern within a theme, these disconfirming cases were retained and reported rather than omitted, to avoid overstating the uniformity of the findings.

### Dependability

It was supported through the maintenance of an audit trail documenting coding decisions, data management steps, and analytic memos throughout the research process, allowing the analytic process to be traced and reviewed. Peer debriefing was conducted with the research supervisor/a co-investigator, who reviewed and challenged emerging codes and themes at key points during analysis.

### Confirmability

It was supported through the same audit trail and peer debriefing process, which allowed interpretations to be checked against the raw data rather than resting solely on the researcher’s individual judgement. Analytic memos documented the reasoning behind coding and thematic decisions, including instances where later analysis prompted revision of earlier interpretations, providing a transparent record of how findings were derived from the data.

### Transferability

It was supported through the provision of detailed contextual information about the study setting, participant characteristics, and data collection procedures, allowing readers to assess the applicability of the findings to other settings or populations. However, as this study was conducted among women receiving treatment at a single tertiary referral facility in Ghana, using a small purposive sample, the findings are not intended to be statistically generalizable and should be interpreted within this context.

## Results

### Sociodemographic characteristics of HIV-positive postpartum mothers

Table 1 summarises the socio-demographic and clinical characteristics of participants. The majority were widowed, with only one participant currently married. Educational attainment was generally modest, with most participants having secondary-level education or below, and one participant with no formal education. Disease stage was similarly distributed between Stage II (five participants) and Stage III (three participants), and treatment duration varied considerably, from 2 to 12 months.

**Table 1:** Socio-demographic and clinical characteristics of participants.

| Participant | Marital status | Education | Disease stage | Treatment received | Duration on treatment (months) |
| --- | --- | --- | --- | --- | --- |
| A | Widowed | None | Stage III | Surgery, RT, CT | 12 |
| B | Single | Secondary | Stage II | Chemotherapy | 6 |
| C | Single | Secondary | Stage II | CT, RT | 6 |
| D | Widowed | Secondary | Stage III | Chemotherapy | 8 |
| E | Widowed | Secondary | Stage III | Radiotherapy | 2 |
| F | Single | Tertiary | Stage II | Chemotherapy | 3 |
| G | Married | Tertiary | Stage II | Surgery, RT | 6 |
| H | Divorced | Secondary | Stage II | CT, Surgery | 6 |

### Thematic findings

Table 2 presents an analysis of the eight interviews, generating five overarching themes: disclosure as a gatekeeping process, reluctance to seek support despite need, consolidation of networks around a single anchor, social support as a precondition for treatment continuity, and disruption of participants’ caregiving identities. Each theme comprised several subthemes grounded in participants’ accounts.

**Table 2:**
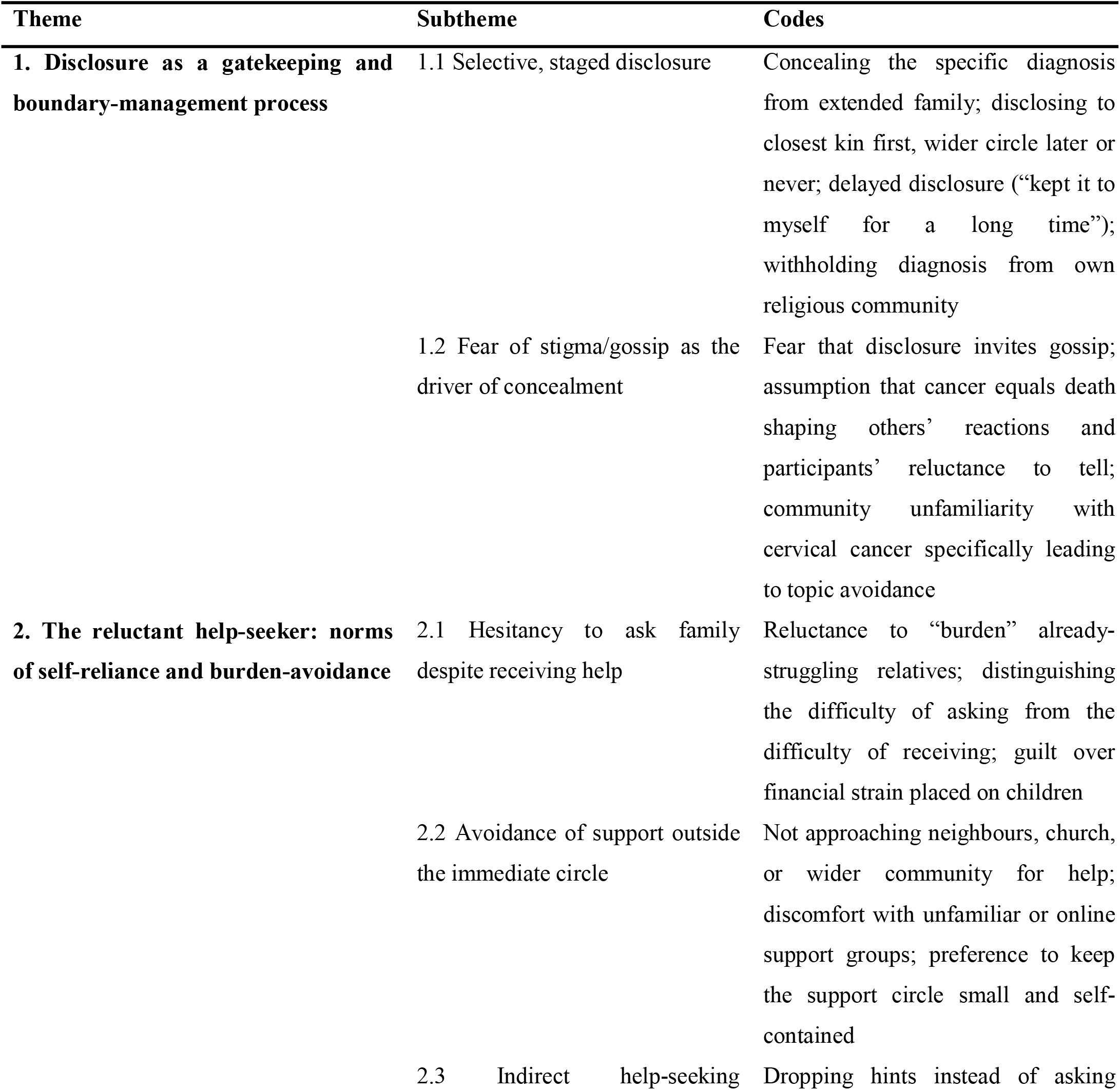
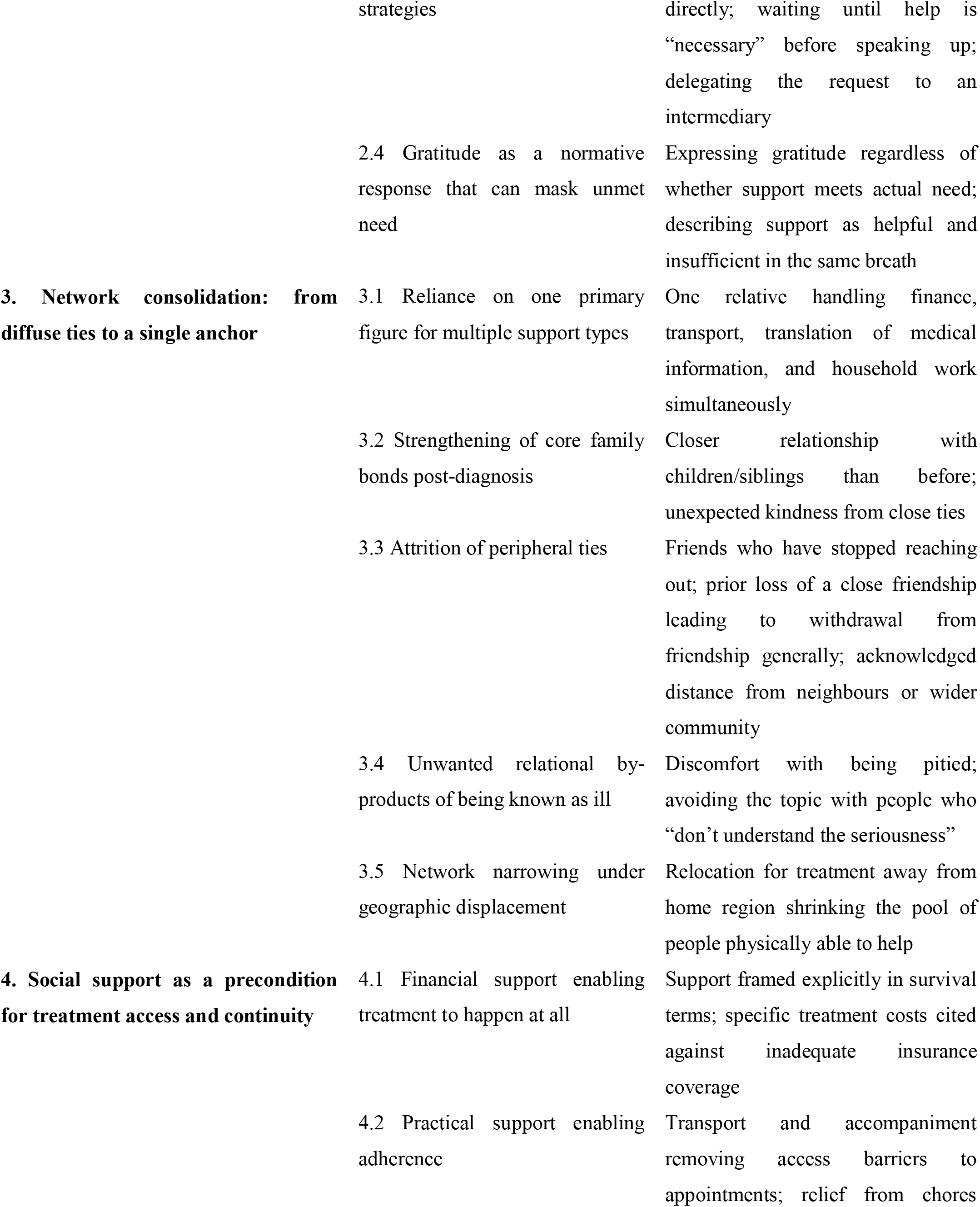
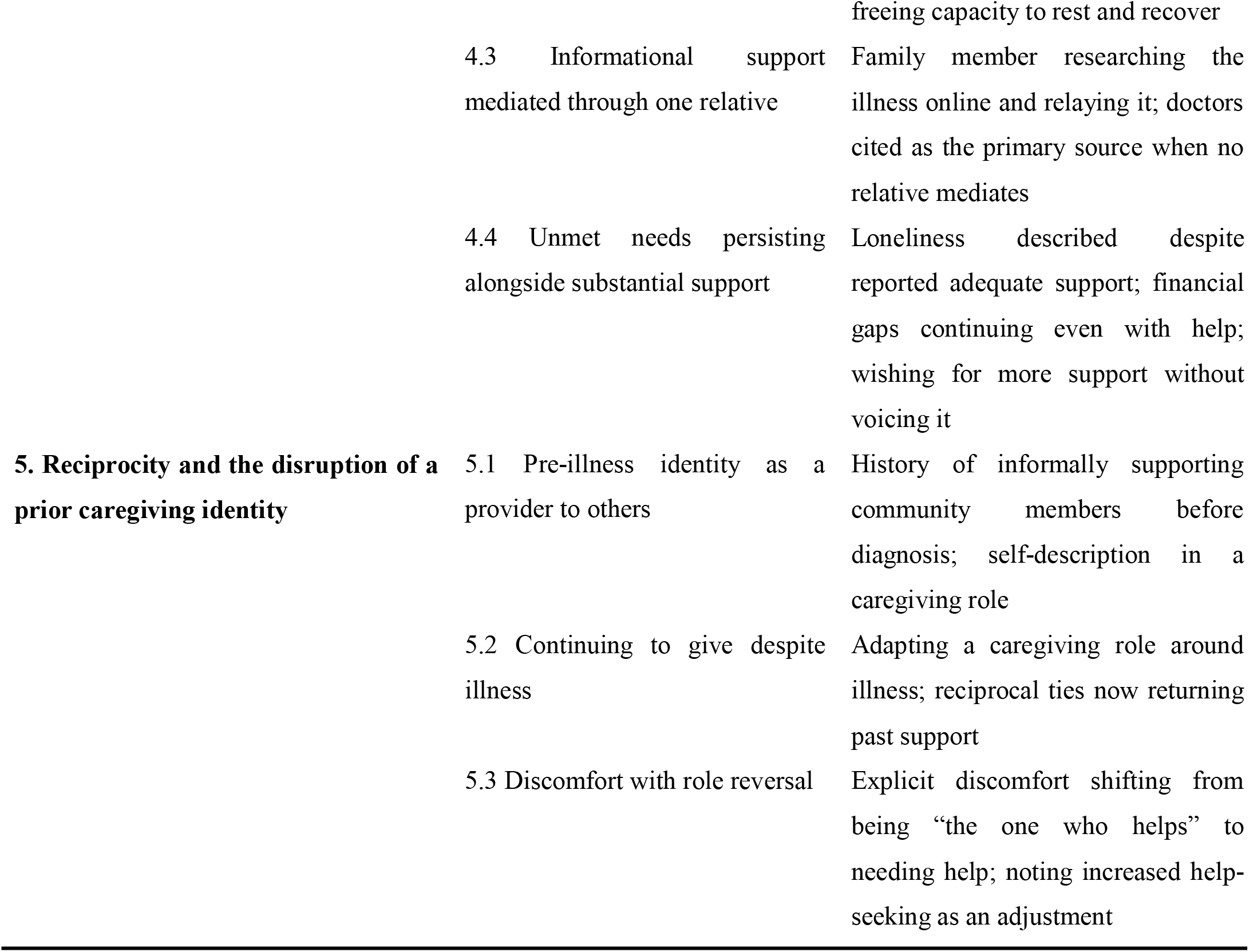
Thematic findings.

### Disclosure as a gatekeeping and boundary-management process

Across the dataset, disclosure of the cervical cancer diagnosis emerged as a controlled, deliberate process rather than a spontaneous act. Participants managed carefully who was told, what was told, and when, effectively using disclosure as a mechanism for determining who could enter their circle of support and who could not. This management operated at two levels: the sequencing and completeness of what was shared with different relational circles, and the underlying anxieties about stigma and misunderstanding that drove this caution. Read together, these subthemes suggest that a woman’s support network during cervical cancer treatment is not a fixed structure inherited from her existing relationships, but one actively shaped, in real time, by her own disclosure decisions.

### Selective, staged disclosure

Disclosure of the diagnosis was rarely a single event. Several participants withheld the specific diagnosis from extended relatives while disclosing only that they were generally unwell, and where fuller disclosure occurred, it tended to follow a deliberate sequence beginning with the closest relational ties before extending, if at all, to a wider circle.

> *“None of them (extended relatives) know the specific disease. If I tell them, they’ll tell everyone. They only know I’m unwell.”* (Participant A)
>
> *“It took me some time to tell them all the details of my illness in the beginning. I kept it to myself for a long time.”* (Participant C)
>
> *“I told the family members closest to me first. I also told a few friends.”* (Participant G)

### Fear of stigma and gossip as the driver of concealment

Underlying this selective disclosure was an anticipation of stigma and a perception that a cervical cancer diagnosis would be misunderstood, gossiped about, or treated as a death sentence, which participants linked directly to their decisions to conceal or delay disclosure.

> *“If I tell them, they’ll tell everyone.”* (Participant A)
>
> *“Most people think when you get cancer, then you’ll surely die from it.”* (Participant C)
>
> *“Most of them didn’t know about cancer of the cervix. Or maybe they don’t care. So I avoid the topic with some of them.”* (Participant G)

### The reluctant help-seeker: norms of self-reliance and burden-avoidance

Despite universally describing themselves as recipients of meaningful support, participants displayed a consistent and somewhat paradoxical reluctance to actively seek that support. This reluctance manifested across four related patterns: hesitancy to ask family members directly for help, avoidance of support from outside the immediate family circle, indirect strategies for signalling need without making an explicit request, and a normative expression of gratitude that could obscure the true extent of unmet need. Taken together, these patterns complicate any straightforward reading of support as simply “adequate” or “inadequate”: what participants received appears to reflect not only what was available to them, but also what they were willing to ask for.

### Hesitancy to ask family despite receiving help

Although every participant described receiving family support, most also described marked reluctance to request it, out of concern for relatives they perceived as already financially stretched.

> *“I feel like I’m putting too much pressure on my kids because they have their own financial struggles.”* (Participant B)
>
> *“The only difficulty is in asking, not in receiving.”* (Participant D)
>
> *“I try to limit asking them for help…I still feel I’m asking for too much.”* (Participant E)

### Avoidance of support outside the immediate circle

Beyond the immediate family, participants consistently described avoiding requests for help from neighbours, congregations, or unfamiliar support groups, preferring to keep the circle of support small and self-contained.

> *“I don’t ask outsiders for help. Even my church doesn’t know…I don’t want to burden others too.”* (Participant C)
>
> *“I haven’t really reached out to other groups, mainly because I’m not that close with a lot of people.”* (Participant B)
>
> *“I don’t get that much help from outsiders.”* (Participant G)

### Indirect help-seeking strategies

Where a need for support did exist, some participants avoided direct requests altogether, instead delaying, hinting, or waiting for support to be offered rather than asked for.

> *“I usually wait until it’s necessary before I say anything. Sometimes I’ll drop hints or mention the struggles I’m facing, hoping that someone will offer to help without me directly asking.”* (Participant E)
>
> *“I don’t like to ask for help unless it’s necessary.”* (Participant G)

### Gratitude as a normative response that can mask unmet need

A related pattern was the consistent expression of gratitude regardless of whether support was described as sufficient, which in several accounts appeared to function as a social obligation that could obscure the extent of unmet need when support was assessed.

> *“When you’re struggling to get by…and someone gives you something small, you can’t say add more. You have to take it like that and show gratitude.”* (Participant A)
>
> *“I’m pleased with the support they give me. I’m really grateful. I always show gratitude.”* (Participant D)
>
> *“It’s not sufficient, but they’re also taking care of their own families.”* (Participant F)

### Network consolidation: from diffuse ties to a single anchor

Rather than drawing on a broad and evenly distributed network, participants’ support structures consistently narrowed around one or two central figures, while relationships at the periphery of the network receded or, in some cases, disappeared altogether. This consolidation took several related forms: dependence on a single anchor for multiple types of support simultaneously, a strengthening of core family bonds following diagnosis, an attrition of more peripheral relationships, unwanted relational costs such as being pitied, and a further narrowing of the available network where participants had relocated for treatment. This pattern was not universal; two participants explicitly reported no change to their networks, but where it did occur, it suggests that the demands of cervical cancer treatment function as a sorting mechanism, revealing which relationships are resilient enough to sustain caregiving over time.

### Reliance on one primary figure for multiple support types

Rather than describing a broad, evenly distributed network, most participants relied on one individual, typically a daughter, son, or sibling, who carried the majority of the practical, financial, and informational support burden.

> *“My daughter comes with me all the time…she was still the one to accompany me.”* (Participant A)
>
> *“My daughter is the one who handles most of that.”* (Participant E)
>
> *“My daughter comes with me most of the time. So she’s the one who talks to the staff.”* (Participant H)

### Strengthening of core family bonds post-diagnosis

Several participants described their closest family relationships strengthening following diagnosis. This pattern was not universal: two participants (D, H) explicitly reported no change in their relationships, which is retained here as a disconfirming case rather than treated as noise.

> *“My bond with my family has become better than before, especially with my children and my sister.”* (Participant G)
>
> *“My relationship with my siblings and children has grown stronger because they have supported me in ways I didn’t expect.”* (Participant E)
>
> *“I became closer with some people who showed me kindness when they heard I wasn’t feeling too well, which was unexpected.”* (Participant C)

### Attrition of peripheral ties

Alongside the strengthening of core ties, more peripheral relationships were consistently described as distant, uninvolved, or receding.

> *“I’ve also stopped hearing from a few friends who don’t really play an important role in my life.”* (Participant G)
>
> *“I’m not that close with my neighbours, so I haven’t gotten much help from them.”* (Participant C)
>
> *“I’m not very close with my current community members.”* (Participant B)

### Unwanted relational byproducts of being known as ill

Being known as unwell also carried unwanted relational costs, particularly the experience of pity, which participants distinguished from the support they valued.

> *“What I’ve noticed is that people feel pity. I don’t really like that.”* (Participant F)
>
> *“Sometimes I feel like people don’t really understand the seriousness of my situation…so I avoid the topic with some of them.”* (Participant G)

### Network narrowing under geographic displacement

For participants who relocated away from their home region to access treatment, the pool of people physically able to provide support narrowed to those who had travelled with them.

> *“Since we came here though (Accra), my daughter has been in charge of cooking, accompanying me and everything else.”* (Participant E)

### Social support as a precondition for treatment access and continuity

Social support was rarely framed by participants as a source of comfort running alongside formal treatment. Instead, it was described in far more consequential terms, as the condition that determined whether treatment could be accessed, afforded, and continued at all. This was evident in how participants discussed the financial stakes of support, the practical logistics it enabled, the informational role it played in helping them navigate treatment decisions, and the needs that remained unmet even where support was substantial. Set against a backdrop of insurance coverage that every participant described as inadequate, this theme positions informal social support not as a supplement to the formal health system, but as something closer to a load-bearing substitute for its gaps.

### Financial support enabling treatment to happen at all

Participants did not describe financial support primarily as a source of comfort, but as a determinant of whether treatment could be accessed and continued at all, against a backdrop of insurance coverage that all eight participants described as inadequate.

> *“I pay more than 4,500 cedis every two weeks…I wouldn’t have been able to pay for my treatment.”* (Participant C)
>
> *“I wouldn’t be able to get treatment at all. Where would I get the money?”* (Participant E)
>
> *“I don’t think I would have survived. I wouldn’t even be able to be in the midst of people.”* (Participant H)

### Practical support enabling adherence

Practical support, particularly transportation and relief from household responsibilities, was consistently linked to participants’ ability to attend appointments and adhere to treatment schedules.

> *“My daughter’s help with transportation means I can get to my appointments on time, and not having to worry about cooking on days when I’m too weak has made it easier for me.”* (Participant A)
>
> *“Having someone to help with meals and chores has made it easier to stick to my treatment schedule.”* (Participant C)
>
> *“She’s also in charge of all the other house chores because we’re away from home. Not having to worry about that helps.”* (Participant E)

### Informational support mediated through one relative

Understanding of treatment and medical information was, for several participants, filtered almost entirely through a single family member who researched or interpreted it on their behalf.

> *“My daughter often helps me understand what the doctors say and what my options are. She also looks up information online and explains it to me.”* (Participant A)
>
> *“My daughter did a lot of research online and shared what she found with me. She talked to the doctors and asked questions I wouldn’t have thought to ask.”* (Participant E)

### Unmet needs persisting alongside substantial support

Despite describing support as helpful, several participants also reported persistent unmet needs, including loneliness and ongoing financial strain, indicating that reported adequacy of support and the presence of unmet need are not mutually exclusive in this dataset.

> *“I’m thankful for the support I receive, but I sometimes wish it were more…it’s hard not to feel a bit lonely sometimes.”* (Participant A)
>
> *“I also wish I had more people to help my kids with funding my treatment.”* (Participant B)

### Reciprocity and the disruption of a prior caregiving identity

For some participants, receiving support during treatment sat uneasily alongside a pre-existing identity as a provider of care and support to others. This tension surfaced in three related ways: participants’ descriptions of their pre-illness role as caregivers within their family or community, their attempts to continue that role, in adapted form, despite illness, and the discomfort they expressed at the reversal of roles that illness had forced upon them. This theme suggests that the psychological experience of receiving support cannot be separated from participants’ prior social identities: for women who saw themselves as habitual givers, becoming a recipient carried a cost distinct from the practical burden of need itself.

### Pre-illness identity as a provider to others

A number of participants described themselves, prior to diagnosis, as habitual providers of support to others in their family or community.

> *“I am like a motherly figure to a lot of my daughter’s friends and even the people in my community…I also raised my little brother, and now he’s the one looking after me.”* (Participant H)
>
> *“When it comes to helping others, I do it a lot…So everyone liked me.”* (Participant A)

### Continuing to give despite illness

Some participants continued this role in adapted form during treatment, and described the people they had previously helped now returning that support.

> *“Even though I can’t teach all the people I was teaching before, I invite some of them to my house so they continue their lessons with me.”* (Participant G)
>
> *“When I was stronger, I also used to help others. And those people have helped me a lot; some still are.”* (Participant D)

### Discomfort with role reversal

For participants accustomed to being the one who helps rather than the one who is helped, needing to rely on others represented a discomforting reversal, distinct from the practical or financial difficulty of receiving support.

> *“I don’t like the way it makes me feel, and I’m used to being the one who takes care of others.”* (Participant G)
>
> *“I just didn’t need as much help before. So I tend to ask more than before.”* (Participant F)

## Discussion

This study set out to examine women’s social networks and the social support exchanged within them during cervical cancer treatment at a tertiary referral facility in Ghana specifically, who becomes involved, how disclosure shapes that involvement, what forms of support are exchanged, and whether women perceive that support as adequate to meet treatment-related needs, including their ability to access and continue treatment itself. The findings show that support networks in this context were neither fixed nor freely accessible: they were actively shaped by women’s own disclosure decisions, constrained by a reluctance to ask for help even where need was evident, and consistently narrowed around a single primary caregiver rather than distributed across a wider network. Beyond documenting these patterns, the study’s central contribution is to show that, for these women, social support functioned less as a comfort accompanying treatment and more as a precondition for whether treatment could be accessed and continued at all. In this sense, the study does not primarily re-establish that cervical cancer is a heavy burden in Ghana, a fact already well established epidemiologically, but explains how women live through treatment within that burden.

Disclosure emerged in this study as an active, managed process through which women determined who could enter their circle of support, rather than a passive by-product of diagnosis. Selective and staged disclosure telling closest kin first, withholding the specific diagnosis from extended relatives, and in some cases concealing the illness from religious communities altogether functioned less as secrecy for its own sake and more as a deliberate strategy for controlling exposure to anticipated judgment. This interpretation is consistent with qualitative work from South Africa, where women receiving curative cervical cancer treatment described experiencing misinformation, stigma, and hesitancy to disclose their illness, often originating from family members and even medical professionals [20]. A related pattern has been documented among Turkish women with cervical cancer, who described experiences of social judgment and isolation alongside spiritual coping strategies that sustained hope and resilience, with stigma limiting support-seeking for some women even as social support alleviated loneliness for others [21].

This qualitative pattern is reinforced by quantitative evidence from Zambia, where higher levels of family and friend support were independently associated with significantly lower odds of stigma among cervical cancer patients [22]. Read alongside the present findings, this suggests disclosure and support are not sequential but mutually reinforcing: women who disclose selectively to trusted ties may, in effect, be securing the very support associated with lower stigma, while withholding disclosure from wider circles limits exposure to the judgment that erodes it.

At the same time, the present findings extend this literature in one respect. Where Williams et al. [20] and Ayşin et al. [21] primarily documented disclosure hesitancy as a psychological or emotional burden, women in the present study described disclosure explicitly as a mechanism of boundary control a way of determining, in practical terms, who would be drawn into the demanding work of providing support during treatment, and who would not. This reframes selective disclosure not simply as concealment driven by shame, but as an active strategy for managing the size and composition of one’s support network under conditions of scarce time, energy, and trust. Gabonese women’s accounts of the psychological repercussions of cervical or breast cancer disclosure similarly describe the diagnosis as reinforcing feelings of stigma, isolation, and disruption to social relationships [23], lending some support to this boundary-management interpretation, though that study’s focus on psychological repercussions rather than network composition means the parallel is suggestive rather than a direct confirmation.

A striking feature of this dataset is that reluctance to seek support was as prominent in women’s accounts as the support itself. Women consistently described hesitating to ask family members directly for help, avoiding requests to neighbours, congregations, or unfamiliar support groups, and, where a need did arise, signalling it indirectly through hints rather than explicit requests. This pattern resonates with supportive care needs research from Nepal, where cervical cancer patients reported psychological burdens that included explicitly feeling a “burden to partner/family,” alongside financial, physical, and informational needs [24], evidence that the reluctance to seek support documented in the present study is layered onto genuine, pre-existing need rather than an absence of it.

The present findings add a further layer: participants did not simply under-report need passively, but actively managed how support was requested, delaying appeals until “necessary” or relying on an intermediary to make requests on their behalf. This suggests burden-avoidance in this context operates less as a fixed personality trait and more as a socially calibrated strategy. Structural quantitative evidence from Ethiopia offers a partial explanation for why this calibration matters so much in practice: supportive care needs among breast and cervical cancer patients at a tertiary centre were found to be prevalent across nearly all patients and every assessed domain, with unmet need significantly greater among patients without health insurance [25], meaning the informal, relationally negotiated support women in the present study were reluctant to request was, for many, filling a gap the formal system had not closed.

A further pattern complicates a purely deficit-based reading of this reluctance: several participants paired expressions of gratitude with descriptions of support they simultaneously characterised as insufficient, suggesting gratitude functioned as a social obligation rather than a straightforward indicator of satisfaction. This carries methodological implications beyond the present study, where research relies on self-reported adequacy of support as an outcome measure without qualitative depth to unpack how gratitude norms shape those reports; sufficiency may be systematically overstated.

Rather than describing broad networks of support, participants consistently relied on a single relative, most often a daughter, son, or sibling who carried the bulk of the practical, financial, and informational support required throughout treatment. A small qualitative case study from Indonesia similarly found that family members functioned as primary anchors, providing continuous presence and financial support that directly sustained patients’ treatment motivation [26], lending direct support to the centrality and concentration of family support observed in the present study.

This concentration carries clear costs for the anchor figure that this study, by design, could not capture directly, since only women receiving treatment were interviewed. Complementary evidence from Tanzania offers insight into this gap: male partners caring for women with cervical cancer described psychosocial distress, economic burden, and altered social responsibilities in their caregiving role [27]. Read alongside the present findings, this suggests the apparent strength of a single-anchor support arrangement should not be mistaken for a sustainable one; it may instead reflect a redistribution of burden rather than its resolution.

Alongside this consolidation, participants described a broader restructuring of their networks: closer bonds with immediate family, alongside a fading of more peripheral relationships. Two participants departed from this pattern, reporting no discernible change following diagnosis; a disconfirming case was retained here rather than smoothed over, cautioning against treating network restructuring as a universal response to diagnosis.

Perhaps the most consequential finding of this study is that participants did not describe social support as supplementary to their medical care, but as determinative of whether that care could be accessed and sustained at all. Similar disruption has been documented elsewhere in the region: women with cervical cancer in Ethiopia described financial difficulty and disrupted social relationships as central features of their treatment and follow-up experience [28], and Ethiopian women more broadly have described cervical cancer as a burden that extends beyond the individual to affect families financially and psychosocially, with support from family and religious practice serving as the primary coping mechanisms available to them [29]. Work examining barriers to retention in cervical cancer care in Uganda similarly found that financial constraints, lack of partner support, and inadequate information, rather than clinical factors, were the dominant drivers of disengagement from care, with social support identified as a key facilitator of continued engagement [30].

Read together with these findings, the present study’s contribution is to make visible the specific mechanism by which informal support offsets these structural gaps not through a single, generalized form of assistance, but through a combination of financial contribution, transportation, household relief, and informational mediation, often concentrated in one individual, as established above. Intervention research offers a proof of concept for what formalizing this support could achieve: a pilot support programme for South African women undergoing radiotherapy produced statistically significant improvements across most measured support domains following implementation [31], suggesting the benefits currently generated informally by a single anchor figure could, in principle, be extended more equitably through structured programmes.

Despite the centrality of support to treatment continuation, participants did not describe their needs as fully met. Loneliness and ongoing financial strain persisted for several women alongside support they otherwise valued. This coexistence of substantial support and persistent unmet need is not contradictory; rather, it suggests even well-functioning informal support arrangements have a ceiling, beyond which family and community networks alone cannot fully substitute for adequately resourced formal care systems.

A theme that extended beyond the study’s original focus on the receipt of support was the discomfort several participants expressed at needing to rely on others, having previously understood themselves as providers of care within their own families and communities. This finding was not anticipated by the study’s stated objectives, which centred on the networks and support women received rather than their own prior caregiving roles; its emergence illustrates a recognised strength of qualitative descriptive research: its capacity to surface participant-driven concerns beyond a study’s original framework. Comparable psychosocial dimensions of cervical cancer have been documented in the United States, where stigma, empowerment, physical and psychological effects, and social support emerged as interconnected themes among Black, Latina, and Chinese American survivors, with social support types nuancedly associated with symptom burden [32], indicating that the psychological weight of illness in this population is shaped by more than the practical availability of help.

The present findings suggest a further, more specific dynamic: for women accustomed to occupying a caregiving role, the practical necessity of receiving help during treatment appears to carry an identity cost distinct from the logistical burden of need itself. This specific mechanism role reversal as a source of psychological discomfort independent of unmet practical need does not appear to have been directly examined in the existing cervical cancer literature reviewed for this study, suggesting it may represent a genuine, if narrow, contribution of the present findings rather than a confirmation of prior work.

These findings carry direct implications for cervical cancer care in Ghana and similar settings. Where informal, family-based support substitutes for elements of formal care that the health system does not yet reliably provide transportation, financial assistance, and help navigating treatment information facilities have an opportunity to formalize and extend what currently depends on the availability of a single family member. Evidence from patient navigation programmes elsewhere supports the feasibility of this approach: a scoping review found patient navigation was associated with improved screening uptake, reduced time to diagnostic resolution, and improved treatment engagement across the cancer care continuum [33], while the pilot intervention discussed above demonstrated that structured support can produce measurable improvements even within a short implementation period [31]. Given this study’s finding that support was, for several women, a precondition for accessing treatment at all, strengthening formal patient navigation and financial-support mechanisms should be considered not as an enhancement to quality of care but as a determinant of treatment access itself, with direct relevance to Sustainable Development Goal 3 (Good Health and Well-Being), specifically Target 3.4 (reducing premature mortality from non-communicable diseases, including cancer) and Target 3.8 (universal health coverage encompassing access to essential health services without financial hardship).

The finding that disclosure functions as a gatekeeping mechanism, shaped by anticipated stigma, further suggests community-level stigma reduction and cervical cancer awareness efforts remain necessary alongside clinical interventions; women cannot draw on support they are unwilling to seek. This has relevance to Sustainable Development Goal 5 (Gender Equality), given that stigma surrounding a reproductive-health-associated cancer disproportionately affects women’s willingness to seek social and institutional support, and to Sustainable Development Goal 10 (Reduced Inequalities), insofar as the burden of unformalized, family-dependent support is likely to fall hardest on women with the smallest or most financially constrained networks.

Finally, the disproportionate and, at times, distressing burden placed on single anchor caregivers, consistent with existing evidence from male partner caregivers in the region [27], points to a need for caregiver-inclusive models of cancer support rather than models that implicitly treat the patient as the sole unit of concern, consistent with a more comprehensive interpretation of SDG 3’s aim of ensuring healthy lives and well-being for all, extended here to include those who sustain the health-seeking of others.

## Conclusion

This study shows that social networks and social support during cervical cancer treatment in Ghana are not fixed or freely available resources, but ones actively constructed through selective disclosure, constrained by a persistent reluctance to ask for help, and typically concentrated around a single family anchor rather than distributed across a wider circle. For the women in this study, that support functioned as more than a source of comfort: it shaped whether treatment could be accessed and sustained at all, extending the significance of informal support well beyond its usual framing as a supplement to formal cancer care. These findings are drawn from eight women’s accounts at a single tertiary referral facility and should be understood as offering depth and insight into this particular context rather than results intended to generalize numerically beyond it. Future qualitative research should examine these dynamics from the perspective of the anchor caregivers themselves, whose experiences this study could not capture directly, and longitudinal qualitative work following women across the full course of treatment would help clarify how disclosure decisions, network composition, and support needs evolve rather than at a single point of contact. Extending this line of inquiry to women managing cervical cancer without an identifiable primary caregiver would also help establish whether the access and continuity challenges identified here are more acute in the absence of a consolidated support network.

## Strengths and limitations

This study has several strengths. Data collection followed established practices for enhancing credibility, including expert and pilot review of the interview guide, verification of transcripts against original recordings and field notes, and verbatim citation of participants’ own language throughout analysis rather than reliance on paraphrase. Disconfirming cases, most notably the two participants who reported no change in their social relationships following diagnosis, were deliberately retained and reported rather than omitted, guarding against an overstated or artificially uniform account of the findings. An audit trail of coding decisions and analytic memos was maintained throughout, and peer debriefing was used to challenge emerging codes and themes at key points in the analysis. The use of a purposive, criterion-based sampling strategy, continued until thematic saturation was judged to have been reached, supports the depth and relevance of the accounts obtained.

The study also has limitations that should be considered when interpreting the findings. Data were collected from women themselves; the perspectives of the family members who provided support, particularly the primary anchor figures identified across several themes, were not captured, and their experiences of caregiving during this period remain unknown from this study alone. The study was conducted among eight women at a single tertiary referral facility in Accra, and while saturation was judged to have been reached within this sample, the findings reflect the experiences of women who accessed care at this particular facility and should not be assumed to transfer uniformly to women treated in different facility types, disease stages, or regions of Ghana. Of the fifteen women initially approached, seven did not participate, including two who declined specifically to preserve the privacy of their diagnosis; it is possible that women managing the most restrictive disclosure boundaries are underrepresented in this sample, which would tend to understate rather than overstate the significance of the disclosure-related findings. As with all self-report qualitative data, accounts of support adequacy may have been shaped by social desirability and by the gratitude norms identified in the findings themselves, which may have led some women to describe support in more favourable terms than their circumstances otherwise suggested.

## List of abbreviations

COREQ: Consolidated Criteria for Reporting Qualitative Research
GCP: Good Clinical Practices
HPV: Human papillomavirus
IDIs: In-depth Interviews

## Declarations

### Ethics approval and consent to participate

Ethical approval for this study was obtained from the University of Health and Allied Sciences Research Ethics Committee (UHAS-REC A.4[042]23-24) and from the management of Korle-Bu Teaching Hospital (KBTH-ADM/000297/2024) prior to data collection. The study adhered to the ethical principles outlined in the Declaration of Helsinki [34] and the principles of Good Clinical Practice (GCP). All participants were fully informed, both verbally and in writing, about the study’s purpose, procedures, potential benefits, and risks before being invited to participate. Written informed consent was obtained from each participant to ensure voluntary participation and to affirm her right to withdraw at any stage without consequence. Permission to audio-record interviews was obtained as part of the consent process. Confidentiality and privacy were maintained throughout data collection, transcription, and analysis, with study data securely stored and accessible only to the research team. Participants received no financial or material compensation for their involvement in the study.

## Consent for publication

Not applicable.

## Availability of data and materials

The datasets used and/or analysed during the current study are available from the corresponding author on reasonable request.

## Author contributions

AA and FR conceived the study. AA supervised proposal development and data collection. AA, DKD, FR, and RED led data analysis and interpretation and drafted the initial manuscript. AA, DKD, FR, RED, and DA provided technical input, critically revised the manuscript for important intellectual content, approved the final version, and accept responsibility for the integrity of the work.

## Data Availability

All data produced in the present study are available upon reasonable request to the authors

## Acknowledgements

The authors thank the staff and administrators of the National Centre for Radiotherapy, Oncology and Nuclear Medicine, Korle-Bu Teaching Hospital, for their support, and the women who shared their experiences with the research team.

